# Post-operative patterns of failure and outcomes reveal opportunities to optimize treatment of recurrent pancreatic cancer

**DOI:** 10.1101/2025.11.08.25339815

**Authors:** John Michael P. Tomagan, Ahmed M. Elamir, Peter Q. Leung, Salwan Al Mutar, Syed M. Ali Kazmi, Matthew R. Porembka, Nina N. Sanford, Patricio M. Polanco, Shahed N. Badiyan, Robert D. Timmerman, Herbert J. Zeh, Todd A. Aguilera

## Abstract

**Background/Objectives:** Recurrent pancreatic adenocarcinoma (PDAC) after curative resection is challenging to treat. We characterized recurrence patterns and outcomes, focusing on frequency of single-organ liver or lung metastases, and their distribution of oligometastases (1-5 lesions) versus polymetastases (> 5 lesions).

**Methods:** Patients were evaluated after curative resection, and recurrence was classified as local, distant, or combined. Patients with metachronous disease and who received treatment and had single organ lung or liver disease were identified as MetachPDAC, followed by OligoMetach (1-5 lesions) and PolyMetach (> 5 lesions). Demographic features, disease free survival, and overall survival (OS) from surgery and time of recurrence were assessed using the Kaplan-Meier method.

**Results:** Of 572 patients who underwent surgery for pancreas cancer, 366 (64%) recurred: 90 (25%) local, 195 (53%) distant, and 81 (22%) combined. Median OS was shortest for patients who experienced combined failures (19.3 mo) versus those with local (34.8 mo) or distant failures (26.6 mo; p<0.0001). In patients with MetachPDAC recurrence with single-organ metastases (n=112), oligometastatic or OligoMetach (n=85, 76%) was more common as 35% of all distant metachronous recurrence and associated with longer OS (29.9 vs 24.3 mo; p=0.0089). Among OligoMetach, liver metastases (n=61, 72%) were more frequent, but lung only metastases (n=24, 28%) was associated with superior OS (63.8 vs 25.5 mo; p=0.0001). In the OligoMetach subgroup, patients receiving radiotherapy with chemotherapy (n=20, 24%) had longer OS than chemotherapy alone (68.4 vs 26.2 mo; p=0.0002).

**Conclusions:** Oligometastatic MetachPDAC is associated with longer survival than PolyMetach, represents 35% of metachronous failures, and radiotherapy may improve outcomes within the subset. These results highlight the value of recurrence pattern–based treatment strategies, though confirmation in larger prospective studies is needed.

## INTRODUCTION

Pancreatic ductal adenocarcinoma (PDAC) continues to be one of the leading causes of cancer-related deaths despite therapeutic advancements in recent years (*1, 2*). A significant number of patients either present with metastatic disease or develop distant metastases after diagnosis. After surgical resection of the primary cancer a significant proportion of patients develop a recurrence with metastatic disease. In the ESPAC-4 trial evaluating chemotherapy after surgery, over 65% of patients developed recurrent disease and 60% of those recurrences were with distant metastatic disease (*3*). Even as we transition into a neoadjuvant (NA) era with the PREOPANC studies among others, meta-analyseses show recurrence rates of 70% with slightly lower rate of 63% after NA therapy compared to 74% for upfront surgery (*4*).

Recurrent disease may occur locally, at distant sites, or as a combination of both. While local failure can range from 35% to 60%, distant relapse–most often in the liver– is more common ranging in up to 80 to 90% of patients (*5, 6*). Importantly, patients with isolated local recurrences often live longer than those with distant spread, suggesting additional local therapy, such as radiotherapy, may offer benefits in some cases (*7–9*). These observations underscore the critical need to better understand recurrence patterns and develop tailored salvage strategies to improve quality of life or extend survival.

Among patients with distant recurrence, prognosis is shaped by both the site and burden of metastases. The concept of oligometastatic disease, typically defined as 1-5 metastatic lesions, has emerged as a potential window for curative-intent therapy across multiple cancers, including PDAC (*10–14*). Distinguishing oligometastatic from polymetastatic (>5 tumors) PDAC may be critical for treatment planning, as Elamir et al. reported poor survival of only 5 months in patients with PDAC following polyprogression whenever it was observed (*13*). Metachronous pancreatic adenocarcinoma (MetachPDAC)–distant recurrence ≥ 6 months after initial diagnosis per ESTRO/EORTC consensus-present unique management challenges (*15*). While systemic therapy remains the standard for metastatic PDAC, several studies suggest that patients with metachronous pulmonary metastases who undergo lung resection experience improved survival compared to those managed non-surgically (*16, 17*). These findings raise the possibility that select patients with metachronous, single-organ, oligometastatic pancreas cancer may benefit from local ablative radiotherapy in addition to systemic treatment. Given the heterogeneity of recurrence, further investigation is needed to refine treatment strategies for this subset.

There is a critical gap in knowledge of how to classify disease after recurrence and how to select treatment strategies beyond the next like of systemic therapy. Here we present our single institution patterns of failure after surgical resection. We hypothesized that patients with MetachPDAC with limited (1-5 lesions) and single-organ metastases (OligoMetach) have improved survival outcomes and may benefit from more intensive local therapies. Here we show that OligoMetach consists of up to 35% of all recurrent disease and that radiotherapy may help improve survival in this population. Accordingly, formal recognition of this population and development of personalized treatment strategies represent critical next steps to extend survival and refine precision oncology for PDAC.

## METHODS

### Study Design

IRB of University of Texas Southwestern gave ethical approval of this work. UTSW IRB, STU072018-037.

### Inclusion and Exclusion Criteria, and Study Cohorts

This study included all patients diagnosed with PDAC who had resection and who were treated at our institution between 2007 and 2023. The inclusion criteria were as follows: patients with localized and curable disease stage (I-III by the American Joint Committee on Cancer) who underwent curative surgical resection, or those who were seen at our institution following resection at an outside facility, patients who had a histopathological diagnosis of PDAC and patients who developed disease recurrence (local, distant, or both) after surgery. We focused on MetachPDAC, patients who developed distant failure ≥ 6 months after the primary diagnosis as per ESTRO and EORTC consensus (*15*), further classified MetachPDAC as those with single organ liver or lung disease (excluding patients with peritoneal metastasis or other metastatic sites), who received treatment. The exclusion criteria were patients with two primaries, patients with neuroendocrine tumors of the pancreas and patients with synchronous metastases, defined as developing distant failure at or within 6 months of the primary diagnosis. While we focus on patients with single-organ MetachPDAC, all patients with distant recurrences regardless of metastatic site, were included in the analysis to provide comprehensive understanding of recurrence patterns in this population.

The MetachPDAC cohort was subdivided into two groups based on the number of metastases, 1-OligoMetach cohort who developed up to five metastatic lesions, 2-PolyMetach cohort for patients with six or more lesions.

### Outcomes and endpoints

The primary outcome was overall survival, with secondary outcomes being disease-free survival, distant failure-free survival, and survival post-distant failure. We identified survival endpoints as; overall survival from the date of surgery to the date of the last follow-up or death. Disease-free survival was defined from the date of surgery to the date of development of recurrence (local, distant or both). Distant failure-free survival was defined from the date of surgery to the date of distant metastases, and survival post-distant failure was defined from the date of developing metastases to the date of the last follow-up or death.

### Statistical Methods

Descriptive statistics were utilized to summarize demographic variables, presenting means, medians, standard deviations, or proportions as appropriate. Proportions were testing using Fisher’s exact tests. One-way ANOVA was used to compare means of three or more groups. T tests were assumed with unequal variances (Welch). Survival analyses were conducted using the Kaplan-Meier method with log-rank test for overall survival, disease-free survival, distant failure-free survival, and survival post-distant failure. Statistical analysis was done in R studio version 2023.12.0.369 4.3.1 and GraphPad Prism 10.6.0. Hazard ratios and corresponding 95% confidence intervals were reported to provide a robust understanding of survival outcomes.

## RESULTS

### Patient Cohort

Of the 572 patients at our institution who underwent surgical resection for PDAC, 366 (64%) experienced disease recurrence. These patients comprise the recurrent cohort, summarized in **Table 1**, which presents demographic, treatment, and pathologic characteristics by pattern of failure (local, distant, or combined). The median age at diagnosis within this cohort was 65 years. Primary tumors were primarily located in the head and neck of the pancreas (n=270), while fewer were found in the body and tail (n=96). Among patients with recurrent disease, 12.3% received neoadjuvant radiation and 21.3% received adjuvant radiation. Chemotherapy regimens varied within the cohort, though most patients received FOLFIRINOX. Baseline characteristics were comparable across failure groups, except for sex and tumor site.

**Table 1.** Baseline Patient Characteristics. local vs. distant vs. combined failures

| Characteristic | Local failure, N = 90<sup>1</sup> | Distant failure, N = 195<sup>1</sup> | Local and distant failure, N = 81<sup>1</sup> | P<sup>2</sup> |
| --- | --- | --- | --- | --- |
| **Age at diagnosis<sup>3</sup>** |  |  |  | >.9 |
| Mean (SD) | 65 (9.7) | 65 (9.6) | 65 (10.3) |  |
| **Sex** |  |  |  | .026 |
| Male | 49 / 90 (54%) | 87 / 195 (45%) | 50 / 81 (62%) |  |
| Female | 41 / 90 (46%) | 108 / 195 (55%) | 31 / 81 (38%) |  |
| **Tumor site** |  |  |  | .022 |
| Head/neck | 75 / 90 (83%) | 142 / 195 (73%) | 52 / 80 (65%) |  |
| Body/Tail | 15 / 90 (17%) | 53 / 195 (27%) | 28 / 80 (35%) |  |
| **Radiation received** |  |  |  | .79 |
| Neoadjuvant | 10 / 90 (11%) | 27 / 195 (14%) | 9 / 81 (11%) |  |
| Adjuvant | 18 / 90 (20%) | 39 / 195 (20%) | 21 / 81 (26%) |  |
| None | 62 / 90 (69%) | 129 / 195 (66%) | 51 / 81 (63%) |  |
| **Neoadjuvant chemotherapy regimen** |  |  |  | .72 |
| No Chemotherapy | 5 / 89.0 (6%) | 16 / 194.0 (8%) | 5 / 81.0 (6%) |  |
| FOLFIRINOX | 42 / 89.0 (47%) | 76 / 194.0 (39%) | 35 / 81.0 (43%) |  |
| Gemcitabine + Abraxane | 8 / 89.0 (9%) | 24 / 194.0 (12%) | 13 / 81.0 (16%) |  |
| Other chemotherapy | 34 / 89.0 (38%) | 78 / 194.0 (40%) | 28 / 81.0 (35%) |  |
| **Margin status** |  |  |  | .70 |
| Involved | 29 / 89 (33%) | 56 / 193 (29%) | 27 / 80 (34%) |  |
| Uninvolved | 60 / 89 (67%) | 137 / 193 (71%) | 53 / 80 (66%) |  |
| **Nodal status** |  |  |  | .159 |
| Positive | 57 / 89 (64%) | 124 / 194 (64%) | 61 / 81 (75%) |  |
| Negative | 32 / 89 (36%) | 70 / 194 (36%) | 20 / 81 (25%) |  |
| **Lymphovascular invasion** |  |  |  | .191 |
| Present | 50 / 86 (58%) | 123 / 185 (66%) | 55 / 77 (71%) |  |
| Not identified | 36 / 86 (42%) | 62 / 185 (34%) | 22 / 77 (29%) |  |

<sup>1</sup> n / N (%)
<sup>2</sup> One-way ANOVA; Fisher's exact test
<sup>3</sup> Representative of 90 local failure, 195 Distant failure, and 81 combined failure patients

From the MetachPDAC cohort, we identified patients with single-organ liver or lung metastases. **Supplemental Table 1** summarizes demographics, tumor location, initial treatment, and postoperative pathologic features, which were similar between the OligoMetach and PolyMetach groups. Of the 366 patients with recurrence, 112 (31%) met criteria for the MetachPDAC cohort. Within this cohort, radiology reports and imaging reviews further distinguished patients with single-organ OligoMetach (n=85, 23% total) and PolyMetach (n=27, 7% of total) (Supplemental Table 1/Figure 1A).

**Figure 1.**
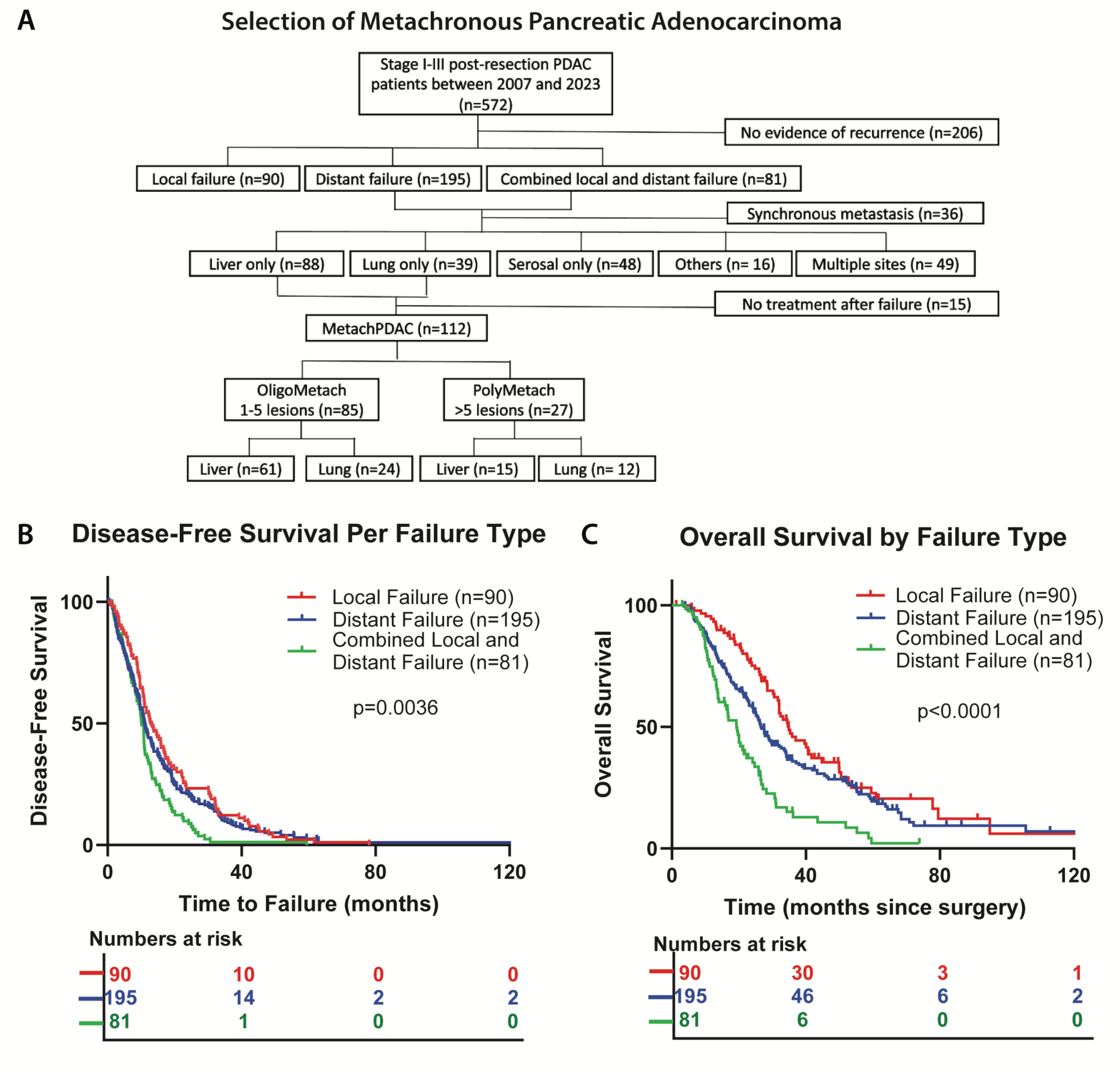
Patterns of recurrence and survival following resection of PDAC. A. Distribution of recurrence among 366 patients with failure, classified as local (n=90), distant (n=195), or combined local and distant (n=81), with further stratification into single-organ MetachPDAC and oligometastatic versus polymetastatic subgroups. B. Disease-free survival (DFS) by recurrence pattern, with longest DFS in local failure compared with distant or combined failure (p=0.0036). C. Overall survival (OS) by recurrence pattern, showing superior outcomes for local failure (34.8 months) versus distant (26.6 months) or combined failure (19.3 months; p<0.0001). All p-values from log rank test.

Among patients with OligoMetach, most primary tumors arose in the pancreatic head/neck (n=61), with fewer in the body/tail (n=24). In the PolyMetach group, 24 tumors were in the head/neck, and 3 in the body/tail. Of those that received radiation, 16 patients (19%) with OligoMetach received neoadjuvant therapy and 17 (20%) adjuvant therapy for total of 39% having had radiotherapy. In the PolyMetach group, 3 (11%) and 10 (37%) patients received neoadjuvant and adjuvant respectively, for a total of 48% having had radiotherapy. FOLFIRINOX was the most common chemotherapy regimen, administered to 49.4% of patients with OligoMetach and 29.6% with PolyMetach. Pathologic features differed between groups: in the OligoMetach cohort, 62.4% had nodal involvement 24.7% positive margins, and 68.2% lymphovascular invasion, whereas in the PolyMetach cohort, these rates were 81%, 41%, and 74% respectively.

### Disease Recurrence and Outcomes Post-Resection

Of the 366 (64%) patients that experienced recurrence, 90 (25%) had local failure, 195 patients (53%) had distant failure, and 81 patients (22%) had combined local and distant failure. **Figure 1A** details the classification of patients regarding local and distant failure, the site of failure, and if they were classified as developing single organ MetachPDAC and OligoMetach or PolyMetach.

The disease-free survival was longest for patients experiencing local failure (13.0 months) compared with distant failure (11.0 months) and combined failure (10.1 months; p=0.0036; **Figure 1B**). Median overall survival was also longest for local failure alone at 34.8 as compared to distant failure (26.6 months) and combined failure (19.3 months; p<0.0001) (**Figure 1C**).

Among patients with metachronous disease, the most common site of recurrence was the liver (n=88, 37%), followed by peritoneal/serosal surface (n=48, 20%) and the lung (n=39, 16%). Less frequent sites included nonregional lymph nodes, adrenal glands, bone, and abdominal wall (n=16, 7%). Multiple metastatic sites were observed in 49 patients (20%) (**Figure 2A**). Survival outcomes differed by site of metastasis. Among the 240 patients with metachronous recurrence, 127 (53%) had single-organ MetachPDAC confined to the liver or lung. When compared with patients who had metastases to other sites, no significant survival differences were observed (p=0.2480) (**Figure 2B**).

**Figure 2.**
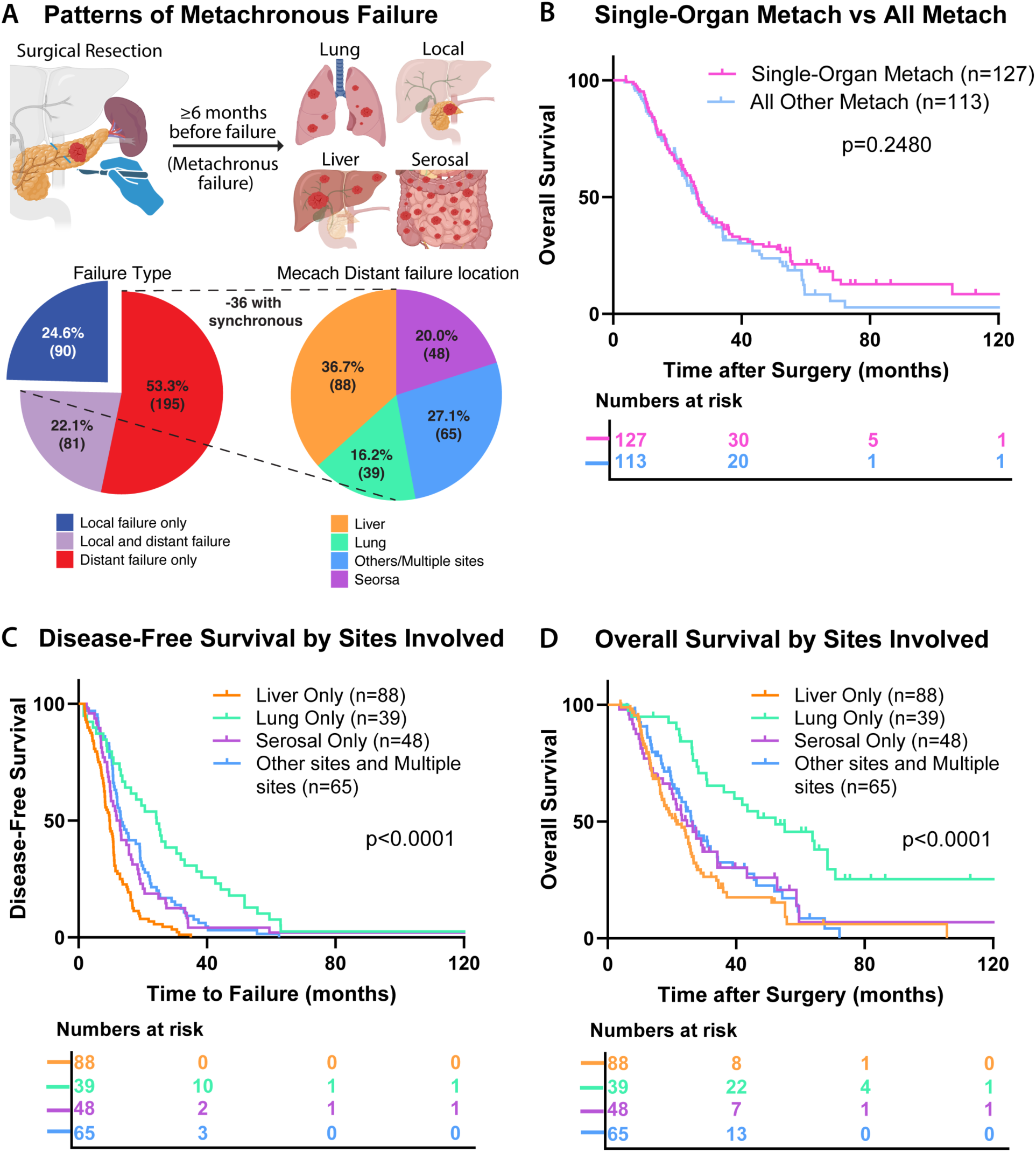
Patterns of recurrence among MetachPDAC and associated survival outcomes. A. Patterns of recurrence among patients with metachronous PDAC, with distant failure as the most common type, followed local and combined. Among those with distant failures, liver was the most common site, followed by peritoneum/serosa, lung, and other less frequent sites. B. Kaplan–Meier curves of OS comparing single-organ liver or lung MetachPDAC to other distant sites, demonstrating no significant difference in OS by recurrence location. C. Kaplan–Meier curves of DFS by metastatic site, showing significantly longer DFS for patients with lung-only recurrences compared with liver, serosal, or other/multiple sites. D. Kaplan–Meier curves of OS by metastatic site, revealing a marked survival advantage in patients with lung-only metastases compared with all other subgroups. All stats by log-rank test.

In terms of outcomes by site of recurrence (liver, lung, serosal, or other/multiple sites), disease-free survival was significantly longer for patients with lung-only metastases (24.3 months) compared with liver-only (9.7 months), serosal-only (12.4 months), and other/multiple sites (13.3 months; p<0.0001) (**Figure 2C**). Similarly, patients with lung-only metastases demonstrated a significantly longer overall survival (52.2 months) compared with serosal-only (24.7 months), liver-only (21.5 months), and other/multiple sites (26.5 months; p<0.0001) **Figure 2D**). These data suggest that based on site of metastases, those with lung only have improved outcome compared to all others as has been observed previously (*17–21*).

### Prevalence and outcomes of single-organ metachronous pancreas cancer

Because single-organ lung and liver recurrences can potentially be treated with ablative radiotherapy, we evaluated outcomes based of this single-organ MetachPDAC group based on the number of metastases. Within the 240 patients with MetachPDAC, 112 patients (47%) had single-organ liver or lung recurrences and received treatment (**Figure 3A**). Of these, 85 were classified as oligometastatic, representing 35% of all metachronous failures and 76% of the single-organ MetachPDAC failures. Notably, oligometastases (n=85, 76%) were more common than polymetastases (n=27, 24%).

**Figure 3.**
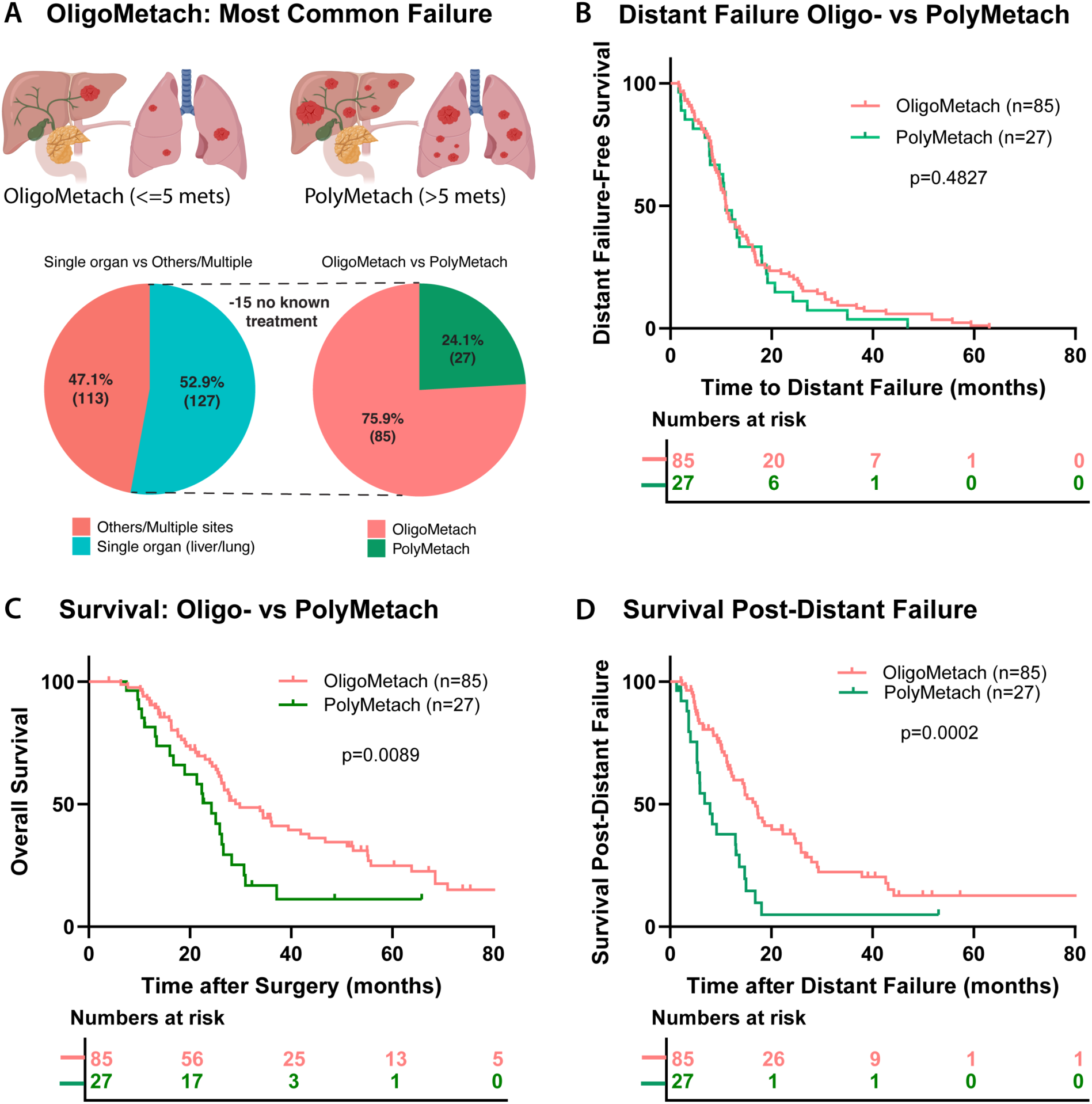
Prevalence and outcomes of oligometastatic versus polymetastatic MetachPDAC. A. Breakdown of patients with metachronous distant recurrence, highlighting the relative frequency of single-organ OligoMetach (≤5 lesions) versus PolyMetach (≥6 lesions). B. Kaplan–Meier curves of distant failure-free survival, showing no significant difference between OligoMetach and PolyMetach patients. C. Kaplan–Meier curves of OS, demonstrating significantly improved survival in OligoMetach compared with PolyMetach patients. D. Kaplan–Meier curves of post-recurrence survival, revealing prolonged survival in OligoMetach patients compared with those with PolyMetach disease. All stats by log-rank test.

We hypothesized that patients with OligoMetach disease would have improved survival outcomes compared with those with PolyMetach disease. While distant failure-free survival from surgery did not differ between groups (p=0.4827; **Figure 3B**), overall survival was significantly longer in the OligoMetach cohort (29.9 months) than in the PolyMetach cohort (24.4 months; p=0.0089; **Figure 3C**). Moreover, at the time of distant failure, patients with OligoMetach demonstrated markedly longer post-failure survival (16.9 vs. 7.9 months; p=0.0002; **Figure 3D**).

### Timing and Survival Between OligoMetach Lung versus Liver Metastases

Given known differences in outcomes between lung and liver metastases, we evaluated whether patients with OligoMetach disease exhibited distinct recurrence timing and survival outcomes. Liver involvement predominated in both subgroups, occurring in 72% of single organ oligometastatic cases (n=61) and 56% of polymetastatic cases (n=15). Lung involvement was less frequent, observed in 28% (n=24) and 44% (n=12) of oligometastatic and polymetastatic cases, respectively (**Figure 4A**). Patients with lung-only OligoMetach experienced significantly longer distant failure–free survival compared to those with liver-only OligoMetach (**15.0 vs. 10.0** months, *p*<0.0001; **Figure 4B**), This delay in recurrence translated into a marked overall survival benefit, with median survival of **40.0** months for lung metastases compared with **20.0** months for liver metastases (*p*<0.0001; **Figure 4C**). Importantly, survival following distant recurrence also favored patients with lung metastases, who demonstrated significantly longer post-recurrence survival than those with liver involvement (*p*=0.0088; **Figure 4D**). Together, these findings highlight two key features of OligoMetach lung metastases—a delayed time to recurrence and prolonged survival after recurrence—suggesting a distinct and more favorable natural history compared to liver metastases.

**Figure 4.**
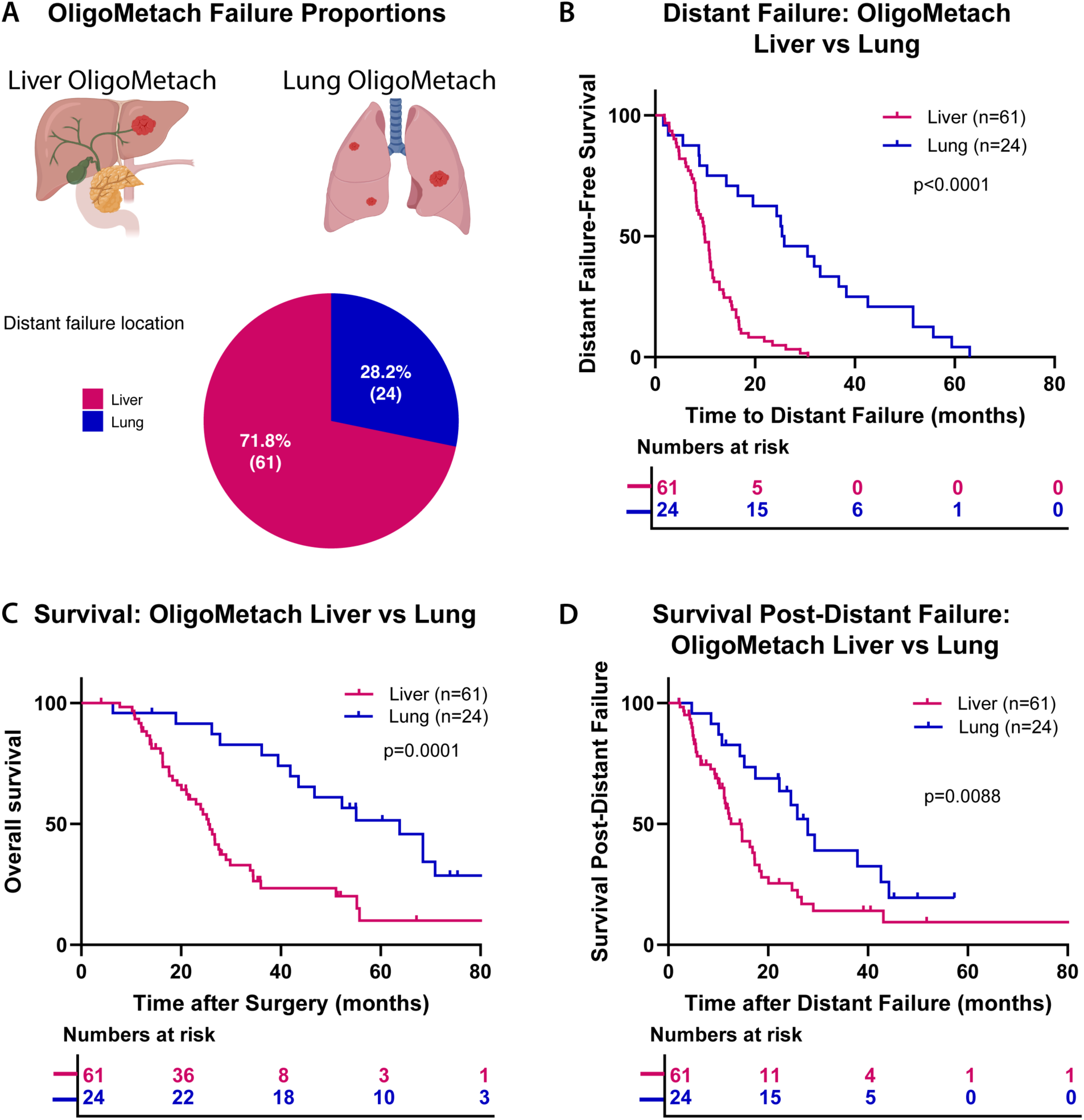
Distinct clinical course of OligoMetach lung versus liver metastases. A. Schematic subgrouping within OligoMetach patients by site of recurrence (lung-only versus liver-only). B. Kaplan–Meier curves of distant failure–free survival, demonstrating significantly later recurrence in lung metastases compared with liver. C. Kaplan–Meier curves of OS, showing superior survival in patients with lung-only metastases compared with those with liver-only disease. D. Kaplan– Meier curves of post-recurrence survival, further highlighting longer survival after recurrence for patients with lung involvement versus liver. All stats by log-rank test.

### Survival after Radiotherapy in those with OligoMetach

Within the OligoMetach subset (n=85), 24% of patients (n=20) received chemotherapy plus radiotherapy and 76% (n=65) received chemotherapy only (**Figure 5A**). Notably, treatment with both therapies was consistently associated with superior outcomes across all endpoints. Patients treated with chemotherapy and radiotherapy had nearly double the distant failure-free survival compared to chemotherapy alone (20.0 vs. 10.3 months, p=0.0015; **Figure 5B**). As anticipated, there was an improvement in overall survival in this group (68.4 vs. 26.2 months; p=0.0002; **Figure 5C**). We also observed that survival after distant failure was significantly longer (37.9 vs. 12.2 months; p=0.0006; **Figure 5D**).

**Figure 5.**
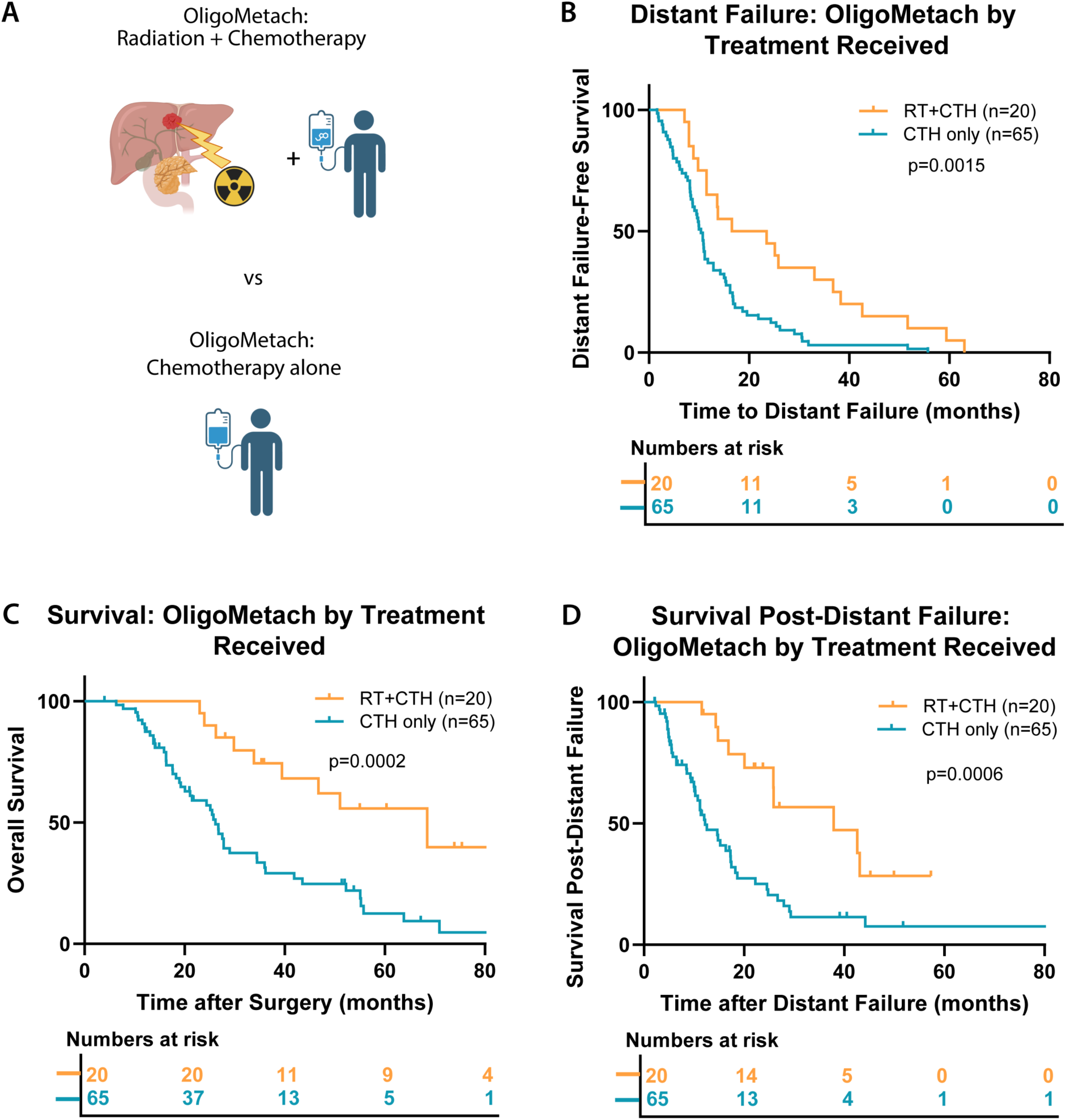
Improved survival outcomes with radiotherapy in OligoMetach PDAC. A. Treatment breakdown of patients with OligoMetach PDAC, stratified by chemotherapy alone versus chemotherapy plus radiotherapy. B. Kaplan–Meier curves of distant failure–free survival, showing a doubling of survival with the addition of radiotherapy. C. Kaplan–Meier curves of OS, demonstrating significantly prolonged survival in patients receiving radiotherapy in addition to chemotherapy. D. Kaplan–Meier curves of post-recurrence survival, revealing longer survival after failure among patients treated with combined modality therapy. All stats by log-rank test.

## DISCUSSION

In this study, we analyzed the patterns of failure and outcomes of 572 at a single institution who underwent resection for PDAC to better understand the timing and patterns of disease. The recurrence rate post-resection was 65%, with local failures observed in 25% of patients, distant failures in 53% and combined local and distant failure in 22%. Patients with local failures had a significantly longer median survival of 34.8 months compared to patients with distant failures (26.6 months) and combined failures (19.3 months). Among patients with distant failures (n=276), 32% had liver-only metastases and 14% had lung only metastases. Patients with lung-only metastases experienced a significantly longer median survival of 52.2 months compared to those with serosal only involvement (24.7 months) and liver only metastases (21.5 months). In the MetachPDAC subgroup, OligoMetach (1-5 lesions) were more prevalent and demonstrated a more durable post-distant failure survival compared to polymetastatic cases. Within the OligoMetach subset, patients receiving radiation and chemotherapy experienced improved survival outcomes compared to chemotherapy alone.(*13, 22*)

Our data align with existing literature on PDAC recurrence patterns, which report high recurrence rates post-surgery, with distant failures being prevalent. A study by Jones et al. reported that 65.6 % of patients experience recurrence after resection, with liver being the most frequent site (*3*). Furthermore, the survival advantage associated with local failures, compared to distant failures, is supported by other studies which suggest local recurrences may be amenable to aggressive local therapies (*7*). Our findings support this observation, as patients with local failure had the most favorable survival outcomes.

While studies investigating oligometastatic pancreatic ductal adenocarcinoma are increasing, a universally accepted definition remains unclear (*11*). Damanakis et al. described the oligometastatic state as involving CA 19-9 levels below 1000 U/mL, four or fewer metastases in the liver or lung, and disease stability or response to first-line chemotherapy in the absence of local consolidation therapy. In a recent systematic review by Leonhardt et al., oligometastatic PDAC encompasses either three or fewer synchronous liver metastases or two or fewer metachronous lung metastases (*11, 12*). In this study, we subdivided the MetachPDAC cohort into two distinct groups based on the number of metastases in a single organ. The OligoMetach cohort included patients with up to five metastatic lesions while the PolyMetach cohort encompassed those with six or more metastatic lesions to refine the distinction between limited metastatic disease and more extensive spread, enabling prognostic evaluation.

Consistent with previously reported series, our study demonstrated that liver metastasis was the most common site of metastasis in both oligometastatic (72%) and polymetastatic (56%) patients and among patients with distant failures (40%). Our findings also highlight the importance of the metastatic site in survival outcomes. Patients with lung-only metastases demonstrated more favorable survival outcomes, with a median survival of 52.2 months compared to 21.5 months for those with liver-only metastases. These findings are similar with studies by Katz et al. and Lovecek et al., which identified the lung as a common site of metastasis among long-term PDAC survivors, with a reported OS of 31.81 months in patients with metachronous pulmonary metastases (*21, 23*). In a similar retrospective study, Liu et al. reported superior median OS for patients with isolated lung metastases (11.8 months) compared to those with isolated liver (6.9 months) or multiple metastases (5.0 months) (*17*). Oweira et al. demonstrated that patients with isolated liver metastatic pancreatic cancer had worse survival outcome compared with those with isolated lung or distant nodal metastases (*24*). These survival differences allude to biological variations in metastatic behavior between organ sites, which may suggest site-specific therapeutic considerations.

Our findings also suggest potential survival benefit of radiotherapy, particularly stereotactic ablative radiotherapy in oligometastatic pancreatic cancer patients compared to chemotherapy alone. Within the oligometastatic subset of patients with metachronous lesions, RT was associated with significantly improved progression-free survival and overall survival. This is consistent with a previous study, where SABR-treated OPanc patients experienced a median polyprogression-free survival of 40 months compared to 14 months in those receiving only chemotherapy (HR = 0.2; 95% CI, 0.07–0.54; P = .0009). Additionally, SABR was associated with OS of 42 months, over double that of the chemotherapy-only cohort (18 months; HR = 0.21; 95% CI, 0.08–0.53; P = .0003) (*13*) Moreover, Scorsetti et al. demonstrated that delivering ablative doses of RT was critical in achieving significant survival benefits, highlighting the importance of treatment intensity among patients with oligometastatic pancreatic cancer (*25*). The EXTEND phase II study also supported the addition of metastasis directed therapy to systemic therapy for patients with oligometastatic PDAC with improvement in PFS in a much more heterogeneous patient population (10.4 months versus 2.5 months; p = 0.030)(*14*). Future prospective studies are warranted to refine patient selection criteria and further validate these findings in the OligoMetach group.

Our study has limitations, including its retrospective design. Biases in patient selection, treatment allocation, and heterogeneity in chemotherapy and radiation treatment approaches could influence survival outcomes. The limited sample size, particularly within the oligometastatic subset, may affect generalizability of our findings. Future research could focus on validating these findings in larger, prospective studies and investigating the underlying mechanisms of the observed outcomes.

In conclusion, this study underscores the role of failure patterns in predicting survival outcomes in recurrent PDAC. Local recurrence is associated with better survival compared to distant failure and combined failures. Patients with OligoMetach exhibit better survival outcomes than patients with single-organ PolyMetach driven by a more durable survival post-distant failure. Therefore, this specific subset of patients may derive benefit from more aggressive metastases-directed local therapies. These findings underscore the need for personalized treatment approaches that consider both the failure pattern and the metastatic burden, which could optimize outcomes in recurrent pancreatic cancer.

## Acknowledgements

We acknowledge the Clinical Data Exchange Network (ClinDEN) team, and the UTSW Tumor Registry team. OpenAI’s ChatGPT large language model (all versions) was used from the date of first release to the submission of the manuscript to confirm proper language, improve conciseness, and enhance clarity. The authors take full responsibility for the integrity of the content. Our patients who put their trust in the multidisciplinary program to personalize treatment options.

## Data Availability

Research data from chart review may be available upon request and appropriate execution of a sharing agreement.

## Supplemental Table

**Table 2.** Baseline Patient Characteristics. Oligometach vs. polymetach

| Characteristic | Oligometach, N = 85<sup>1</sup> | Polymetach, N = 27<sup>1</sup> | P<sup>2</sup> |
| --- | --- | --- | --- |
| **Age at diagnosis<sup>3</sup>** |  |  | .35 |
| Mean (SD) | 65 (9.2) | 63 (9.9) |  |
| **Sex** |  |  | .66 |
| Male | 45 / 85 (53%) | 16 / 27 (59%) |  |
| Female | 40 / 85 (47%) | 11 / 27 (41%) |  |
| **Tumor site** |  |  | .117 |
| Head/neck | 61 / 85 (72%) | 23 / 26 (88%) |  |
| Body/Tail | 24 / 85 (28%) | 3 / 26 (12%) |  |
| **Radiation received** |  |  | .21 |
| Neoadjuvant | 16 / 85 (19%) | 3 / 27 (11%) |  |
| Adjuvant | 17 / 85 (20%) | 10 / 27 (37%) |  |
| None | 52 / 85 (61%) | 14 / 27 (52%) |  |
| **Neoadjuvant chemotherapy regimen** |  |  | .185 |
| No Chemotherapy | 1 / 84.0 (1%) | 1 / 27.0 (4%) |  |
| FOLFIRINOX | 42 / 84.0 (50%) | 8 / 27.0 (30%) |  |
| Gemcitabine + Abraxane | 11 / 84.0 (13%) | 6 / 27.0 (22%) |  |
| Other chemotherapy | 30 / 84.0 (36%) | 12 / 27.0 (44%) |  |
| **Margin status** |  |  | .144 |
| Involved | 21 / 84 (25%) | 11 / 27 (41%) |  |
| Uninvolved | 63 / 84 (75%) | 16 / 27 (59%) |  |
| **Nodal status** |  |  | .099 |
| Positive | 53 / 85 (62%) | 22 / 27 (81%) |  |
| Negative | 32 / 85 (38%) | 5 / 27 (19%) |  |
| **Lymphovascular invasion** |  |  | >.9 |
| Present | 58 / 80 (73%) | 20 / 27 (74%) |  |
| Not identified | 22 / 80 (28%) | 7 / 27 (26%) |  |

<sup>1</sup> n / N (%)
<sup>2</sup> Welch Two Sample t-test; Fisher's exact test
<sup>3</sup> Representative of 85 oligometach and 27 polymetach patients

